# Association between long-term stimulant treatment and the functional brain response to methylphenidate in adolescents and adults with attention-deficit/hyperactivity disorder

**DOI:** 10.1101/2024.12.30.24319766

**Authors:** Zarah van der Pal, Liesbeth Reneman, Henk JMM Mutsaerts, Antonia Kaiser, Marco A Bottelier, Hilde M Geurts, Anouk Schrantee

**Affiliations:** Department of Radiology & Nuclear Medicine, Amsterdam UMC location University of Amsterdam, Amsterdam, Netherlands; Department of Radiology & Nuclear Medicine, Amsterdam UMC location Vrije Universiteit, Amsterdam, Netherlands; CIBM Center for Biomedical Imaging, Lausanne, Switzerland; Animal Imaging and Technology, Ecole Polytechnique Fédérale de Lausanne (EPFL), Lausanne, Switzerland; Accare, Centre for Academic Child and Adolescent Psychiatry, UMC Groningen, Groningen, Netherlands; Dutch Autism and ADHD Research Center, Amsterdam Brain and Cognition, University of Amsterdam, Amsterdam, Netherlands

**Keywords:** attention-deficit/hyperactivity disorder (ADHD), cerebral blood flow, brain development, stimulant medication, pharmacological MRI, neuroimaging

## Abstract

**Background:** Stimulant medication is commonly used by children and adolescents with attention-deficit/hyperactivity disorder (ADHD), however its long-lasting effects on the developing brain remain unclear. In a previous randomized controlled trial (RCT) we found that short-term stimulant treatment influences the functional brain response to an acute methylphenidate-challenge in an age-dependent manner, in line with animal studies suggesting persisting effects on brain development.

**Methods:** In this 4-year naturalistic follow-up of the initial RCT, we investigated the long-term age-dependent effects of stimulant treatment on the functional brain response to methylphenidate in male children and adults with ADHD (n=56; adolescents aged 10-17 years, adults aged 23-43 years). At baseline and 4-year follow-up, we used pharmacological MRI to estimate relative cerebral blood flow (rCBF) before a single-dose methylphenidate-challenge (resting rCBF) and the rCBF-response to a single-dose methylphenidate-challenge. Linear mixed models were constructed to evaluate the effect of stimulant medication use, age and visit on resting rCBF and rCBF-response.

**Results:** We found no evidence for long-term age-dependent effects of stimulant treatment, suggesting that our previously identified short-term effects may be transient. We did identify age-dependent associations between rCBF-response in the medial prefrontal cortex and stimulant treatment, which were already present before treatment initiation but were unrelated to ADHD symptom severity. Moreover, rCBF-response was associated with dopamine D1 receptor distributions in adolescents only.

**Conclusions:** The identified age-dependent associations may potentially be mediated by changes in dopamine- and noradrenaline-related functioning, and may hold predictive value for (extent of) stimulant medication use after ADHD diagnosis in children and adolescents.

## Introduction

Stimulant medication, such as methylphenidate, belongs to the largest class of psychotropic medications prescribed for the treatment of attention-deficit/hyperactivity disorder (ADHD). Methylphenidate blocks the dopamine and noradrenaline transporters in the brain, leading to an increase in extracellular dopamine and noradrenaline [1, 2]. Methylphenidate’s short-term safety has been documented in many studies and its efficacy is among the highest of psychiatric medications. However, despite its widespread and prolonged use, surprisingly little is known on the long-term effects of stimulant medication on the human brain in development.

During adolescence, the brain undergoes rapid development, showcasing specific sensitive periods [3] which render the brain particularly susceptible to interventions, like stimulant medications, which might alter these crucial developmental processes. For example, the ratio of excitatory D1/5 receptors and inhibitory D2/3/4 receptor expression changes throughout development [4-6]. Indeed, animal studies suggest that exposure to stimulant medication at different developmental stages affects the outgrowth of the dopamine system, with long-lasting consequences [7, 8]. As such, administering methylphenidate during crucial developmental periods may imprint enduring effects on brain maturation (‘the neurochemical imprinting hypothesis’ [9, 10]), significantly manifesting as individuals approach adulthood.

In humans, studies investigating the effect of stimulant medication on the development of the dopamine and noradrenaline systems remain sparse, and are hampered by the in-accessibility of these systems without using invasive (i.e. radioactive) imaging tracers. In addition, these studies exhibit a lack of convergent findings [11-15], accompanied by methodological challenges such as the absence of longitudinal treatment study designs. To this purpose, we previously validated the non-invasive method ‘pharmacological MRI’, in which alterations in resting cerebral blood flow (CBF) in response to an acute challenge with methylphenidate are assessed [16-18]. Given the pharmacological targets of methylphenidate, the CBF-response can be interpreted as a proxy for changes in dopamine- and noradrenaline-related brain function. We subsequently applied this technique in a prospective longitudinal randomized controlled trial (RCT) aimed at probing the effects of methylphenidate treatment on brain development (“effects of Psychotropic drugs On the Developing brain” (ePOD) Methylphenidate project, NTR3103). We showed that 4-month treatment with methylphenidate, compared to placebo, decreased the CBF-response to an acute challenge with methylphenidate after one-week washout in stimulant treatment-naive children, but not adults, with ADHD [19]. These findings suggest *short-term* lasting age-dependent effects of methylphenidate treatment on the CBF-response to a methylphenidate-challenge. However, in a related cross-sectional study exploring the *long-term* effects of age-of-first-stimulant treatment on the brain’s response to methylphenidate, we observed no differences in CBF-response between participants that initiated stimulant treatment during childhood and those that initiated stimulant treatment during adulthood, and stimulant treatment-naive participants [20]. However, the group exposed to stimulants during childhood exhibited lower baseline CBF in the anterior cingulate cortex, compared with the group that had not received stimulant treatment. This discrepancy could signify early-induced alterations by stimulant medication on the developing dopamine and noradrenaline systems.

To further investigate the *long-term* effects of stimulant treatment, we here conducted a 4-year naturalistic follow-up of participants from the initial ePOD-methylphenidate RCT, assessing alterations in resting CBF and response to an acute challenge with methylphenidate. We hypothesized that stimulant treatment would induce persistent long-term age-dependent effects on development of the dopamine and noradrenaline systems in the brain. Specifically, we hypothesized that higher stimulant medication use during the naturalistic follow-up would be associated with lower CBF-response in adolescents, but not adults. Moreover, we expected that the age-dependent effects previously identified in our RCT [19] could still be identified after 4 years; in line with the neurochemical imprinting hypothesis which states that neurochemical imprinting effects increase with age in preclinical studies [7, 9, 21].

## Methods

### Study design and participants

The present study constitutes the naturalistic 4-year follow-up assessment of the ePOD-methylphenidate RCT (2016-2019 [22]). The initial RCT was a 16-week double-blind, randomized, placebo-controlled trial of methylphenidate treatment (2011-2015) in stimulant treatment-naive male participants with ADHD [19, 23]. The ePOD-methylphenidate RCT protocol was approved by the Central Committee on Research Involving Human Subjects (identifier NL34509.000.10). The 4-year follow-up assessment was approved by the local medical ethical committee of the Academic Medical Center (NL54972.018.15). All participants and parents or legal representatives of the children provided written informed consent.

In the original RCT, 50 boys (aged 10-12 years) and 49 men (aged 23-30 years) with ADHD were included, based on an a priori power calculation [23]. An experienced psychiatrist diagnosed all participants according to the Diagnostic and Statistical Manual of Mental Disorders (DSM-IV, 4th edition [24]) using a structured interview (Diagnostic Interview Schedule for Children fourth edition, DISC-IV [25]) in children or in parents and the Diagnostic Interview for Adult ADHD [26] in adults (for details on recruitment and exclusion criteria, see Supplementary Materials). For the 4-year naturalistic follow-up assessment, participants were contacted by phone and/or email to ask if they wanted to participate in the follow-up assessment. Exclusion criteria were contraindications to MRI.

The present study reports on the primary outcome of the long-term ePOD-methylphenidate project, namely using pharmacological MRI to assess stimulant medication effects on the CBF-response to an acute challenge with methylphenidate. At baseline and 4-year follow-up, MR scanning was performed before and 90 minutes after a single-dose methylphenidate-challenge (0.5 mg/kg, with a maximum of 20 mg for adolescents and a maximum of 40 mg for adults) (Figure 1A). To exclude possible acute effects of stimulant medication use (half-life: 2-10 hours), participants were asked to stop using stimulant medication one week prior to the follow-up assessment (one-week washout period). A total of 33 adolescents and 25 adults returned for the 4-year follow-up assessment, for details see the results section.

**Figure 1.**
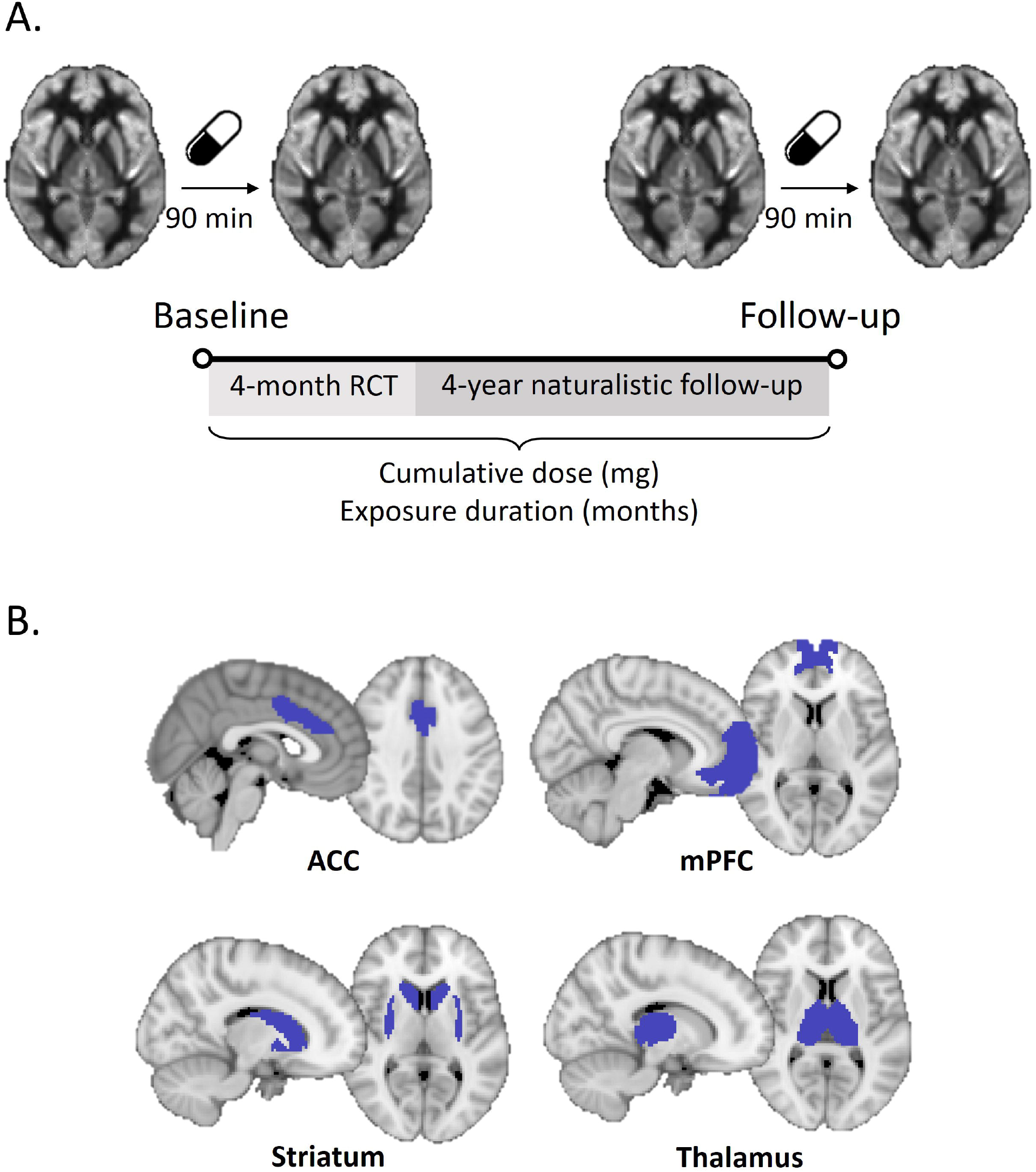
Study design and regions of interest (ROIs). A) At baseline and 4-year follow-up assessment, participants underwent MR scanning before and 90 minutes after a single-dose short-acting methylphenidate-challenge (0.5mg/kg; max. 20 mg for adolescents, max. 40 mg for adults). Stimulant medication use between baseline and follow-up (cumulative dose/exposure duration) was calculated from medication received during the randomized controlled trial (RCT) and pharmacy prescription information from the naturalistic follow-up. B) Predefined ROIs: anterior cingulate cortex (ACC), medial prefrontal cortex (mPFC), striatum and thalamus.

### Stimulant medication use

We calculated stimulant medication use per participant based on medication received during the initial trial and medication prescription information between the trial and 4-year follow-up assessment obtained from participant’s pharmacies. The medication use variables cumulative dose (mg) and exposure duration (months) were extracted. Moreover, we determined age at start of medication use (years), stimulant medication use at follow-up (yes/no) and stimulant treatment-naivety at follow-up (yes/no) (for details, see Supplementary Materials).

### ADHD symptom severity

At baseline and 4-year follow-up, self-reported ADHD symptom scores were assessed using the inattentive and hyperactivity/impulsivity subscales of the Disruptive Behavior Disorder Rating-Scale (DBD-RS [27]) in adolescents, and the ADHD-Rating Scale (ADHD-RS [26]) in adults. Although these scales are primarily used as continuous measures of ADHD symptom severity and are insufficient for establishing a clinical diagnosis, we here include the cut-off values to provide a broader context for interpretation. Clinically significant symptoms were defined as scores >15 on the DBD-RS inattentive and hyperactive/impulsive subscales for adolescents [28] and scores >10 on the ADHD-RS for adults [26].

### MRI - acquisition

MR data were acquired using a 3T MR scanner (baseline: Intera or Achieva, follow-up: Ingenia; Philips Medical Systems, Best, The Netherlands) using an 8-channel receive-only head coil. A pseudo-continuous arterial spin labelling (PCASL) sequence with a gradient-echo echo-planar imaging readout was acquired before (pre-MPH) and after (post-MPH) the methylphenidate-challenge (parameters: TR/TE=4000/14 ms; post-label delay range=1525-2143 ms; label duration=1650 ms; FOV=240×240x119 mm; 75 dynamics; voxel size=3×3×7 mm; 17 contiguous slices; no background suppression). Moreover, we acquired a 3D T1-weighted scan (parameters: TR/TE=9.8/4.6 ms; FOV=256x256x120; voxel size=0.875×0.875×1.2 mm; 120 slices) for registration purposes. Heart rate was measured using a peripheral pulse unit.

### MRI - processing

Image processing and CBF quantification were performed using the ExploreASL pipeline (v1.12.0 Beta [29]) in Matlab (vR2022b). Briefly, the T1-weighted images were segmented into grey matter (GM), white matter and cerebrospinal fluid using the Computational Anatomy Toolbox 12 [30]. Next, for the ASL time series, motion artefacts were detected using motion estimation, and a fixed spike exclusion threshold (1mm) was applied for outlier exclusion. CBF (mL/min/100g) was calculated with the recommended single-compartment model from the perfusion-weighted and M0 images [31]. Since no background suppression was used, the mean of the control images was used as M0 image. Rigid-body registration to the corresponding T1-weighted image was performed for all intermediate and final images, followed by nonlinear registration to MNI space. Finally, visual quality control was performed [29].

For two adolescents, background suppression was used for the ASL sequences at baseline. For four adolescents, at 4-year follow-up, the ASL MR data were exported as perfusion images instead of raw unsubtracted ASL images. Therefore, image processing was adjusted for these six participants (for details, see Supplementary Materials).

Median CBF (mL/min/100g) was estimated for global GM and four predefined regions of interest (ROIs): anterior cingulate cortex (ACC), medial prefrontal cortex (mPFC), striatum and thalamus (Figure 1B; for the Brainnetome parcellation numbers, see Supplementary Table 1). The striatum was selected because it is rich in dopamine transporters, which are targeted by stimulant medication such as methylphenidate. The choice for the ACC, mPFC, and thalamus was based on animal literature demonstrating the largest age-dependent effects of methylphenidate in these important projections of the striatum [8, 10]. Moreover, these four ROIs were assessed in previous short-term studies in the same sample [19, 32].

### Statistical analysis

All analyses were conducted using R v4.4.1 (R Development Core Team, 2011). We calculated CBF for each ROI relative to global CBF (i.e. relative CBF, rCBF), to account for large physiological, vascular, and scanner variability in global CBF (Chen et al., 2011). The rCBF-response to an acute methylphenidate-challenge was defined as *(post-MPH - pre-MPH)/pre-MPH*.

Linear mixed effect models (LMMs; *lme4* package version 1.1-35-5) were used to assess the effect of stimulant medication use (cumulative dose, exposure duration), visit (baseline, follow-up), age group (adolescents, adults) on i) rCBF before a single-dose methylphenidate-challenge (resting rCBF), and ii) the rCBF-response to a single-dose methylphenidate-challenge (rCBF-response). Moreover, we evaluated whether the short-term effects identified in the initial RCT were still present at follow-up using LMMs including the treatment group during the RCT (methylphenidate, placebo) instead of continuous medication use variables as predictors. In all models, we applied multiple comparison correction (FDR=5%) to adjust for the four ROIs evaluated.

The following variables were included as covariates in all models: MR scanner at baseline (Intera/Achieva), standardized scan interval (offset 48 months, corresponding to a 4-year follow-up), and demeaned age at baseline (per age group; to correct for the larger age range at baseline among adults than adolescents). Moreover, mean motion (mm; in the pre-MPH rCBF analysis) and change in mean motion (mm, calculated as *post-MPH - pre-MPH*; in the rCBF-response analysis) were included as covariates in all models. In addition, we explored associations between rCBF, motion and heart rate. Finally, for ROIs showing significant age-dependent medication effects on resting rCBF and rCBF-response, we explored correlations between rCBF and ADHD symptom severity.

### Exploratory whole-brain analyses

To evaluate whether stimulant treatment impacted additional brain regions beyond the four ROIs evaluated in the main analysis, we performed an exploratory whole-brain analysis evaluating the relation between stimulant medication use and (change in) rCBF-response to methylphenidate. In addition, we explored the association between the whole-brain rCBF-response and the spatial distribution of dopamine and noradrenaline receptors to identify which receptor systems may be mediating the observed response (for details, see Supplementary Materials).

## Results

### Participant characteristics

Of the 50 children and 49 adults who participated in the initial trial, 33 children and 25 adults returned for the 4-year follow-up assessment. Baseline characteristics were comparable for participants that returned and did not return for follow-up (Supplementary Table 2). One adolescent and one adult were excluded from analysis due to incomplete MRI data and undisclosed stimulant medication use before baseline, respectively. The final sample consisted of 32 adolescents (aged 11.2±0.9 years at baseline) and 24 adults (aged 29.9±5.0 years at baseline) (Table 1). Post-MPH MRI data was missing for 3 adolescents and 7 MRI data points were excluded from analysis due to insufficient data quality or high motion (>75 excluded frames). This resulted in inclusion of 53 participants at baseline (30 adolescents, 23 adults) and 54 participants at follow-up (30 adolescents, 24 adults) for the resting rCBF analysis, and 50 participants at baseline (27 adolescents, 23 adults) and 52 participants at follow-up (28 adolescents, 24 adults) for the rCBF-response analysis.

**Table 1.**
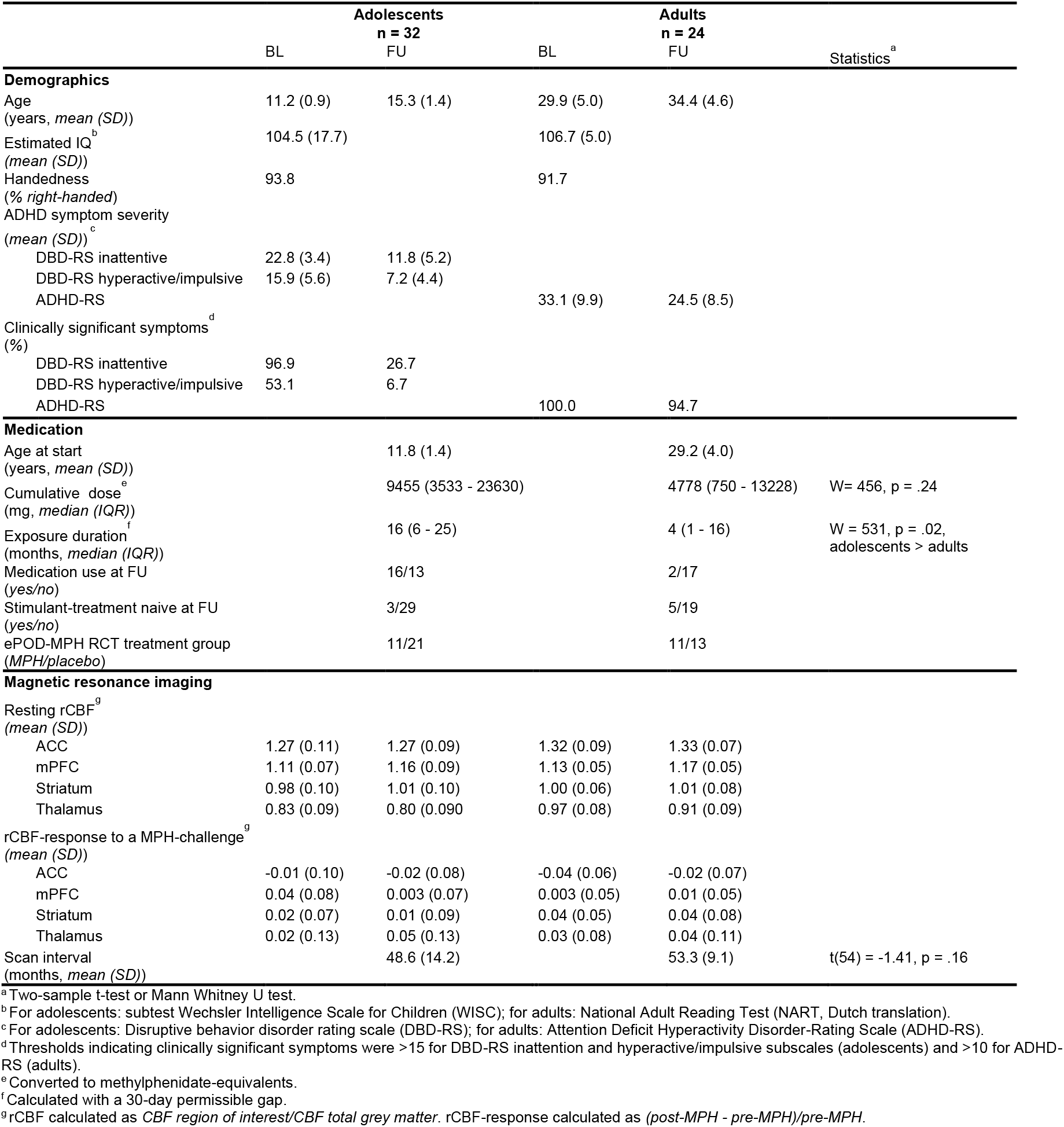
Participant characteristics of the study participants. Data are presented as mean (standard deviation; SD), median (interquartile range; IQR), fraction (yes/no, MPH/placebo) or percentage (%). ADHD=attention-deficit/hyperactivity disorder; BL=baseline; rCBF=relative cerebral blood flow; FU=follow-up; IQ=intelligence quotient; MPH=methylphenidate; RCT=randomized controlled trial.

|  | Adolescents<br>n = 32 |  | Adults<br>n = 24 |  | Statistics<sup>a</sup> |
| --- | --- | --- | --- | --- | --- |
|  | BL | FU | BL | FU |  |
| **Demographics** |  |  |  |  |  |
| Age<br>(years, *mean (SD)*) | 11.2 (0.9) | 15.3 (1.4) | 29.9 (5.0) | 34.4 (4.6) |  |
| Estimated IQ<sup>b</sup><br>(*mean (SD)*) | 104.5 (17.7) |  | 106.7 (5.0) |  |  |
| Handedness<br>(% *right-handed*) | 93.8 |  | 91.7 |  |  |
| ADHD symptom severity<br>(*mean (SD)*)<sup>c</sup> |  |  |  |  |  |
| DBD-RS inattentive | 22.8 (3.4) | 11.8 (5.2) |  |  |  |
| DBD-RS hyperactive/impulsive | 15.9 (5.6) | 7.2 (4.4) |  |  |  |
| ADHD-RS |  |  | 33.1 (9.9) | 24.5 (8.5) |  |
| Clinically significant symptoms<sup>d</sup><br>(%) |  |  |  |  |  |
| DBD-RS inattentive | 96.9 | 26.7 |  |  |  |
| DBD-RS hyperactive/impulsive | 53.1 | 6.7 |  |  |  |
| ADHD-RS |  |  | 100.0 | 94.7 |  |
| **Medication** |  |  |  |  |  |
| Age at start<br>(years, *mean (SD)*) |  | 11.8 (1.4) |  | 29.2 (4.0) |  |
| Cumulative dose<sup>e</sup><br>(mg, *median (IQR)*) |  | 9455 (3533 - 23630) |  | 4778 (750 - 13228) | W= 456, p = .24 |
| Exposure duration<sup>f</sup><br>(months, *median (IQR)*) |  | 16 (6 - 25) |  | 4 (1 - 16) | W = 531, p = .02,<br>adolescents > adults |
| Medication use at FU<br>(*yes/no*) |  | 16/13 |  | 2/17 |  |
| Stimulant-treatment naive at FU<br>(*yes/no*) |  | 3/29 |  | 5/19 |  |
| ePOD-MPH RCT treatment group<br>(*MPH/placebo*) |  | 11/21 |  | 11/13 |  |
| **Magnetic resonance imaging** |  |  |  |  |  |
| Resting rCBF<sup>g</sup><br>(*mean (SD)*) |  |  |  |  |  |
| ACC | 1.27 (0.11) | 1.27 (0.09) | 1.32 (0.09) | 1.33 (0.07) |  |
| mPFC | 1.11 (0.07) | 1.16 (0.09) | 1.13 (0.05) | 1.17 (0.05) |  |
| Striatum | 0.98 (0.10) | 1.01 (0.10) | 1.00 (0.06) | 1.01 (0.08) |  |
| Thalamus | 0.83 (0.09) | 0.80 (0.090) | 0.97 (0.08) | 0.91 (0.09) |  |
| rCBF-response to a MPH-challenge<sup>g</sup><br>(*mean (SD)*) |  |  |  |  |  |
| ACC | -0.01 (0.10) | -0.02 (0.08) | -0.04 (0.06) | -0.02 (0.07) |  |
| mPFC | 0.04 (0.08) | 0.003 (0.07) | 0.003 (0.05) | 0.01 (0.05) |  |
| Striatum | 0.02 (0.07) | 0.01 (0.09) | 0.04 (0.05) | 0.04 (0.08) |  |
| Thalamus | 0.02 (0.13) | 0.05 (0.13) | 0.03 (0.08) | 0.04 (0.11) |  |
| Scan interval<br>(months, *mean (SD)*) |  | 48.6 (14.2) |  | 53.3 (9.1) | t(54) = -1.41, p = .16 |

<sup>a</sup> Two-sample t-test or Mann Whitney U test.
<sup>b</sup> For adolescents: subtest Wechsler Intelligence Scale for Children (WISC); for adults: National Adult Reading Test (NART, Dutch translation).
<sup>c</sup> For adolescents: Disruptive behavior disorder rating scale (DBD-RS); for adults: Attention Deficit Hyperactivity Disorder-Rating Scale (ADHD-RS).
<sup>d</sup> Thresholds indicating clinically significant symptoms were >15 for DBD-RS inattention and hyperactive/impulsive subscales (adolescents) and >10 for ADHD-RS (adults).
<sup>e</sup> Converted to methylphenidate-equivalents.
<sup>f</sup> Calculated with a 30-day permissible gap.
<sup>g</sup> rCBF calculated as *CBF region of interest/total grey matter*. rCBF-response calculated as *(post-MPH - pre-MPH)/pre-MPH*.

Exposure duration to stimulant medication was higher in adolescents compared with adults (W=531, P=.02), whereas cumulative dose did not differ between age groups (W=456, P=.24; Table 1). Two adult participants were prescribed atomoxetine, a non-stimulant medication for treatment of ADHD, in addition to stimulant medication.

### Stimulant treatment was not significantly associated with change in rCBF over time

In contrast to our hypothesis, we found no evidence for an association between stimulant medication use and change in resting rCBF or rCBF-response over time, in either age group or any of the ROIs (*age*medication*visit interaction*; all P>.10; Table 2). The rCBF-response did not change significantly between baseline and follow-up in either age group or any ROI (*age*visit interaction*; all P>.10) (Table 2B).

**Table 2.**
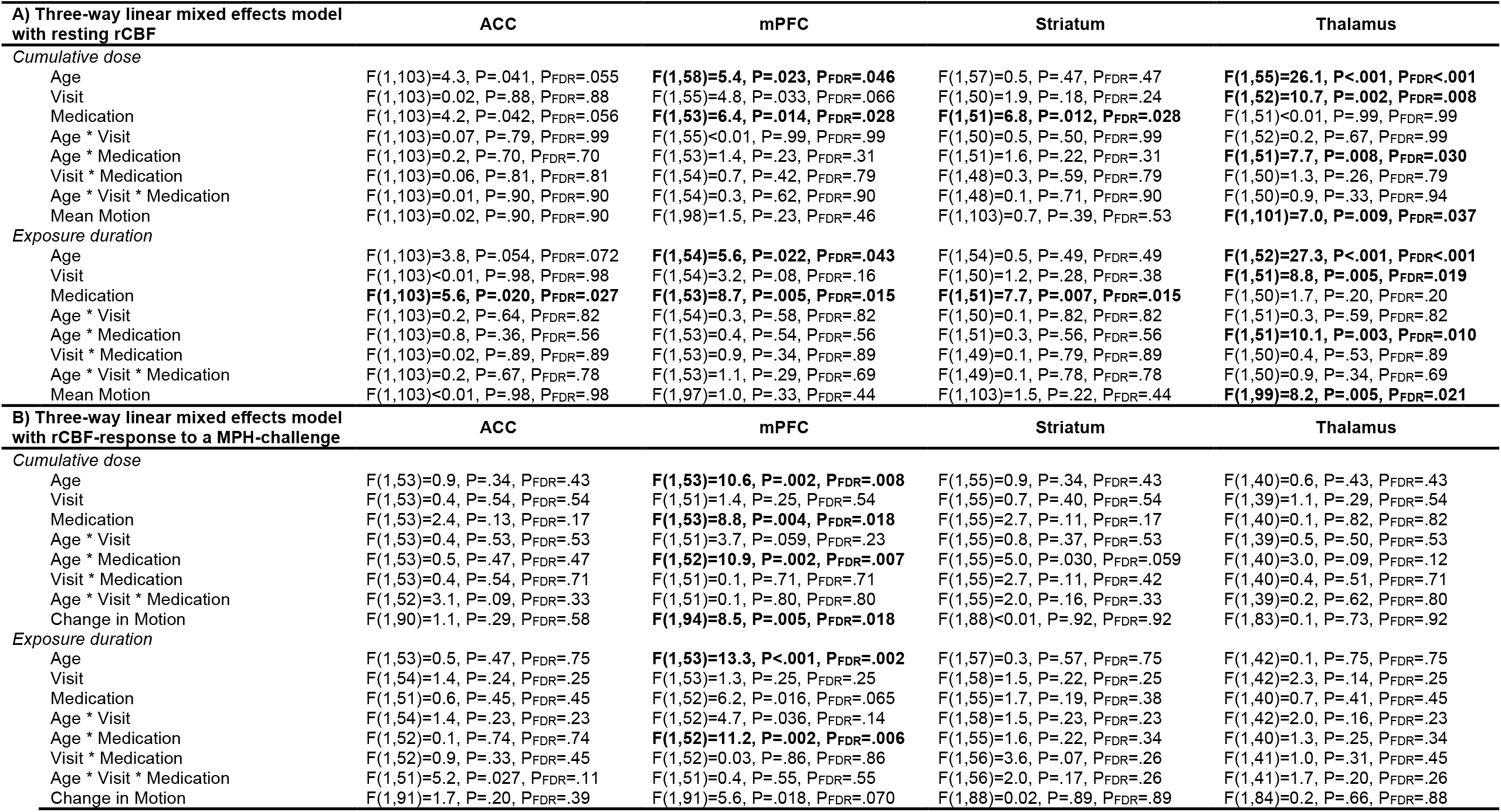
Linear mixed effect model results. Bold values represent significant effects after multiple comparison correction (FDR=5%, 4 comparisons). Separate models were used per medication variable (cumulative dose, exposure duration). The variables mean or change in motion, MR scanner at baseline, baseline age, and scan interval were included as covariates in all models. For readability covariates were excluded from the table, except for motion, which showed a significant effect in some models. ACC=anterior cingulate cortex; rCBF=relative cerebral blood flow; mPFC=medial prefrontal cortex; MPH=methylphenidate; P_FDR_=P-value after FDR multiple comparison correction.

Resting rCBF in the mPFC was consistently higher in adults than adolescents at both visits (*age effect*; all P<.046). Moreover, thalamic resting rCBF showed a decrease between baseline and follow-up, in both age groups (*visit effect*; all P<.019) (Table 2A).

### Extent of stimulant use was significantly associated with resting rCBF and rCBF-response in an age-dependent manner

Across visits, thalamic resting rCBF was significantly associated with stimulant medication use in an age-dependent manner (cumulative dose: F(1,51)=7.7, P=.030; exposure duration: F(1,51)=10.1, P=.010) (*age*medication interaction*; Table 2A). Post-hoc analysis revealed that adults (but not adolescents) taking more stimulant medication, and having a longer exposure duration, showed lower thalamic resting rCBF (Figure 2A). Moreover, across age groups and visits, we found a significant association between stimulant medication use and resting rCBF in the other ROIs (*medication effect*; ACC, mPFC, striatum) (Table 2A), such that participants taking more stimulant medication, and having a longer exposure duration, showed higher resting rCBF.

**Figure 2.**
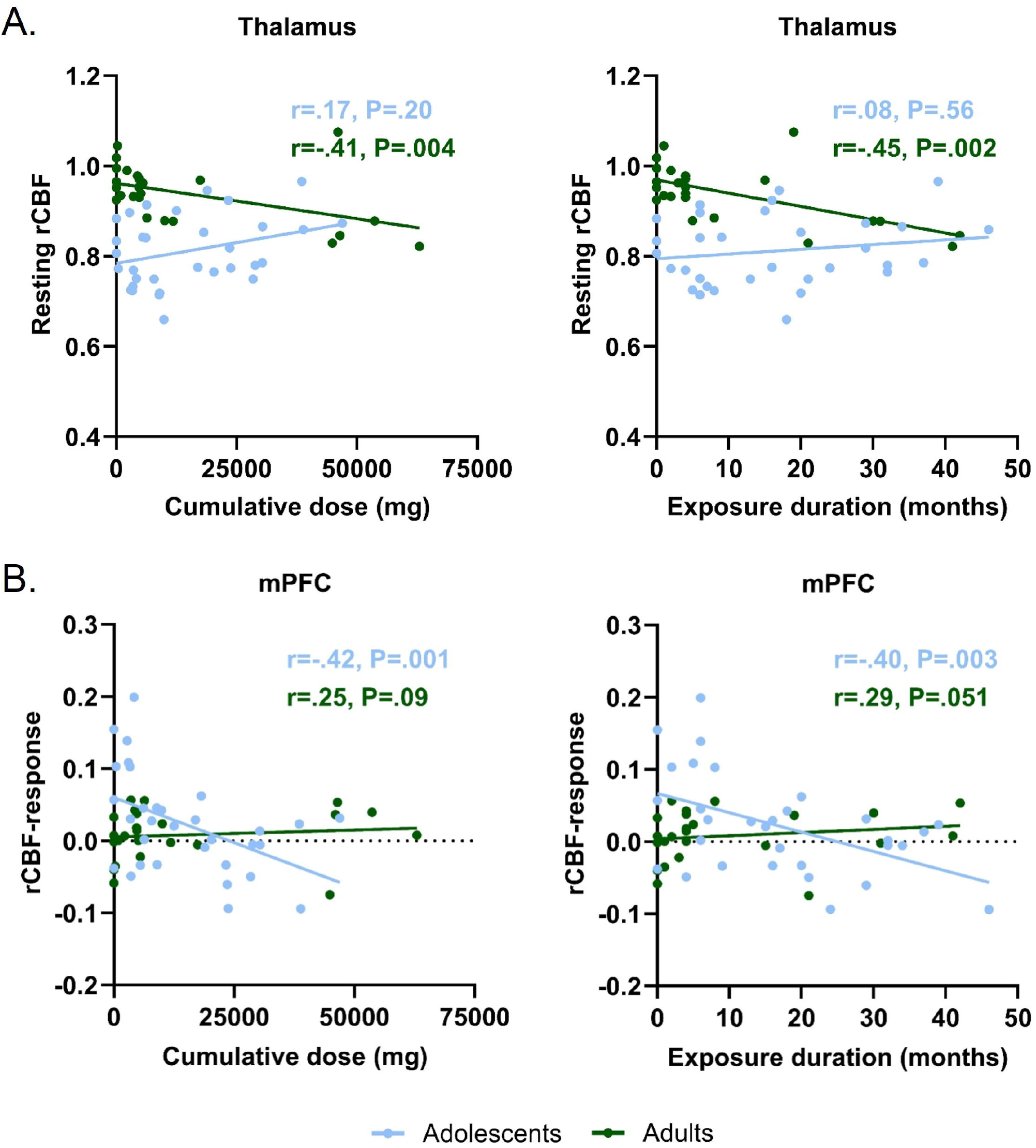
Age*medication interaction effects on resting rCBF and rCBF-response to a methylphenidate-challenge. A) Thalamic resting rCBF was associated with medication use in adults, but not in adolescents. B) mPFC rCBF-response was associated with medication use in adolescents, but not in adults. Data are visualized as the average rCBF values across visits, since the age*medication interaction effects are independent of time. mPFC=medial prefrontal cortex; rCBF=relative cerebral blood flow.

Additionally, rCBF-response in the mPFC was significantly associated with stimulant medication use in an age-dependent manner (cumulative dose: F(1,52)=10.9, P=.007; exposure duration: F(1,52)=11.2, P=.006) (*age*medication interaction*; Table 2B), such that adolescents (but not adults) taking more stimulant medication, and having a longer exposure duration, showed a lower rCBF-response to an acute methylphenidate-challenge (Figure 2B). No such effects were observed in the other ROIs (Table 2B).

### Sensitivity analyses reveal robustness of the findings

We found no evidence that initial treatment allocation during the ePOD-methylphenidate RCT (i.e. methylphenidate or placebo arm) had long-lasting effects on resting rCBF or rCBF-response to a methylphenidate-challenge (Supplementary Table 3). Absolute CBF values were higher in adolescents than adults and reduced between baseline and follow-up in adolescents only (Supplementary Table 4). Moreover, we found no associations between (change in) ADHD symptom severity and (change in) resting rCBF or rCBF-response (Supplementary Table 5; linear Pearson correlations). Heart rate increased after the methylphenidate-challenge, but this was not associated with rCBF-response (Supplementary Tables 6-7). Motion was higher in adolescents than adults, and reduced after a single-dose methylphenidate-challenge in adolescents only (Supplementary Table 8). (Change in) motion was a significant predictor in the thalamic resting rCBF and mPFC rCBF-response models (Table 2), but results were robust with and without motion in these models. We found no associations between rCBF and motion in any of the other models (Supplementary Table 9).

### Whole-brain rCBF-response was significantly associated with spatial distribution of D1 receptors

We conducted an exploratory whole-brain analysis to evaluate whether stimulant treatment impacted additional brain regions beyond the four ROIs evaluated in the main analysis. In both age groups, we found no additional associations between stimulant medication use and (change in) rCBF-response to methylphenidate. Furthermore, exploratory analysis of the association between the whole-brain rCBF-response and the spatial distribution of dopamine and noradrenaline receptors revealed that the rCBF-response maps were significantly associated with D1 receptor distributions in adolescents only. Although the dopamine transporter (DAT), noradrenaline transporter (NAT) and D2/3 receptor are also targets of methylphenidate, we found no evidence for associations of these receptors’ distribution with whole-brain rCBF-response (Table 3).

**Table 3.** Associations between whole-brain rCBF-response to a methylphenidate challenge and dopamine and noradrenaline receptor distributions. Correlation coefficients were obtained using spin tests. Bold values represent significant associations. BL=baseline; rCBF=relative cerebral blood flow; FU=follow-up.

|  | Adolescents |  | Adults |  |
| --- | --- | --- | --- | --- |
|  | BL | FU | BL | FU |
| Dopamine transporter | $r = .12, p = .18$ | $r = .03, p = .75$ | $r = .10, p = .24$ | $r = .04, p = .67$ |
| Dopamine D1 receptor | **$r = .47, p < .001$** | $r = .16, p = .28$ | $r = .16, p = .28$ | $r = .19, p = .21$ |
| Dopamine D2/D3 receptor | $r = .21, p = .15$ | $r = -.05, p = .73$ | $r = .09, p = .59$ | $r = .10, p = .57$ |
| Noradrenaline transporter | $r = -.27, p = .14$ | $r = -.20, p = .31$ | $r = -.27, p = .14$ | $r = -.32, p = .08$ |

## Discussion

In this 4-year naturalistic follow-up study using pharmacological MRI, we found no evidence for long-term age-dependent effects of stimulant treatment on development of resting rCBF and rCBF-response to an acute challenge with methylphenidate in adolescents when compared to adults. This contrasts with our previous short-term findings in the initial RCT [19]. Nevertheless, our findings revealed that thalamic resting rCBF and the mPFC rCBF-response to a methylphenidate-challenge were associated with the extent of stimulant treatment during the 4-year follow-up period in an age-dependent manner. These associations were already present before the commencement of stimulant treatment, but were unrelated to ADHD symptom severity. Finally, we found that rCBF-response was associated with D1 receptor distributions, but not dopamine receptor/transporter distributions.

### No evidence for long-term age-dependent effects of stimulant treatment on development of rCBF

Guided by animal studies and the neurochemical imprinting hypothesis, which suggests that stimulant administration during development has lasting effects that increase with age [7, 9, 21], we hypothesized that the effects we identified in the initial ePOD-methylphenidate RCT [19] would persist long-term. In our previous RCT in the same study population, children receiving 4-month methylphenidate treatment showed reduced CBF responsiveness to a methylphenidate-challenge in the thalamus and striatum, unlike adults or placebo groups. However, our current long-term follow-up data from the same cohort reveal no age-dependent changes over time in CBF-responses, nor dose-dependent stimulant treatment effects.

The discrepancy between our short-term findings in the initial RCT and this long-term follow-up may have several explanations. First, our previously identified short-term effects may be transient due to neuroplasticity - the brain’s ability to adapt and reorganize in response to internal and external stimuli [33]. Second, heterogeneous medication use and treatment adherence during the naturalistic follow-up, in contrast to the RCT, may have resulted in more subtle effects than we could detect [34, 35]. Additionally, the reduced sample size due to participant attrition may have limited our statistical power, meaning that such subtle differences, if present, may have gone undetected. Alternatively, potential stimulant medication effects might become apparent after a longer follow-up period, when the adolescents have reached young adulthood and the neurochemical imprinting effects are suggested to be most pronounced.

Notably, the existing literature presents mixed findings regarding long-term effects of stimulant treatment on brain development. Some studies reported that participants taking stimulant medication were similar to neurotypical controls regarding cortical thickness [36] and brain functioning (at rest [37] or during an attentional task [38]), whereas other studies reported no associations between stimulant treatment and brain development [22, 39, 40] or inconclusive results [15]. Besides methodological heterogeneity and reporting issues, the lack of convergence between these studies may be related to factors like biological factors/genetic predispositions, sample age and behavioral characteristics (for an extensive discussion, see [15, 41]).

### Age-dependent associations between stimulant treatment and rCBF

Exploring the age-dependent associations between rCBF at baseline and subsequent stimulant medication use, we found that this relation was present in adults only for thalamic resting rCBF, and in adolescents only for the mPFC rCBF-response to a methylphenidate-challenge. Notably, these associations remained present also at 4-year follow-up.

Adults with higher thalamic resting rCBF were prescribed less stimulant medication for shorter durations in the years following treatment initiation. That we found this relation solely in adults may be related to the gradual decline in dopamine D2/3 receptors in the thalamus in adulthood [42, 43]. The thalamus plays an important role in attention and cognitive functioning, processes that are commonly affected in ADHD and can effectively be improved with stimulant treatment [38, 44, 45]. Individual differences in thalamic function (e.g. perfusion) may become apparent in adulthood, which may be associated with efficacy of stimulant treatment, and subsequently with the extent of medication use.

Adolescents that exhibited a higher (more positive) rCBF-response to the methylphenidate-challenge at baseline were prescribed less medication for shorter durations, whereas adolescents with lower responsiveness, or even a negative rCBF-response, were prescribed more medication for a longer duration. This finding aligns with the significant maturation of the prefrontal cortex during adolescence, where dopamine D1 and D2 receptor densities peak and then decline into adulthood [5, 46, 47]. The heightened rCBF-response observed in adolescents compared to adults could be attributed to the relatively high density of D1 receptors in the mPFC [6, 48]. Notably, D1 receptors are activated only at higher dopamine levels, and the relatively high dose of methylphenidate used during the challenge may have specifically enhanced D1 receptor activity in the mPFC. This is corroborated by our finding that the whole-brain rCBF-response was associated with D1 receptor distributions in adolescents only. While this whole-brain approach provides valuable neurobiological insights into the mechanisms of methylphenidate action, it does not capture inter-individual variability in receptor distributions. Understanding how receptor distributions vary across individuals—and how this variability influences brain function—could provide crucial insights into the observed differences in medication responses and usage patterns.

Different adolescent neural trajectories may also be associated with varying ADHD phenotypes [49]. In our study, the variation in mPFC rCBF-response was not *linearly* associated with (changes in) self-reported severity of ADHD symptoms. Nonetheless, ADHD symptoms have been shown to follow *non-linear* trajectories across the lifespan; for example, both persistent, remittent and subthreshold symptom trajectories have been described [50, 51]. This may explain the lack of correlation between ADHD symptoms and mPFC rCBF-response in adolescents, but also the differences observed between the adolescent and adult group. Given our study’s design, in which both adolescents and adults were stimulant treatment-naive at baseline, it is possible that individuals diagnosed with ADHD in adulthood may have experienced subthreshold ADHD symptoms during childhood and adolescence, progressing to diagnosis in adulthood, thus constituting an ADHD subpopulation [50]. Moreover, reported distinct genetic variations and risk factors between childhood and adult ADHD diagnoses may contribute to diverse patterns of mPFC function [52].

### Methodological considerations

A critical strength of this study is its longitudinal design with stimulant treatment-naive participants at baseline, ruling out the influence of prior stimulant treatment on brain development. Additionally, the 4-year follow-up period addresses the need to investigate the potentially long-lasting effects of stimulant treatment. Stimulant medication is commonly prescribed to children and adolescents for extended periods; however, neuroimaging studies with long-term follow-up are scarce, and those that do exist often do not include stimulant treatment-naive participants (e.g. [36] and [37]). Furthermore, the use of pharmacological MRI provided insights into how stimulant medication affects brain function over time and presents an interesting technique for subtyping and stratification, contributing to a better understanding of the neural mechanisms that influence treatment patterns.

Our findings should be interpreted in light of several methodological considerations. First, although pharmacy prescription data provides more reliable medication use information during the naturalistic follow-up compared with participant self-report, we assumed complete prescription adherence for all participants (aligning with e.g. [53-55]). However, medication adherence rates can vary substantially [34, 35], necessitating the inclusion of reliable treatment adherence measures in future research [56]. Moreover, while this study focused on males with ADHD [23], future studies should investigate medication effects in females as well [57]. Other limitations include the restricted sample size at follow-up due to attrition and the use of different MR scanners at baseline and follow-up. Still, we were able to identify an age-dependent association between mPFC rCBF-response and stimulant treatment, which may hold predictive value for optimization of treatment approaches for ADHD. Future studies should explore whether long-term effects of stimulant treatment on brain development become evident in young adulthood, when such effects are suggested to be most pronounced [9].

## Conclusion

In conclusion, this study found no evidence for long-lasting effects of stimulant treatment on the functional brain response to methylphenidate. We did identify an age-dependent association between rCBF-response in the mPFC and extent of stimulant treatment, which may hold predictive value for (extent of) stimulant medication use after ADHD diagnosis in children and adolescents. Our findings highlight the value of including stimulant treatment-naive participants and using pharmacological MRI to probe dopamine- and noradrenaline-related brain function. Large-scale longitudinal studies are encouraged to further explore the potential of pharmacological MRI for optimization of treatment approaches in ADHD, and disentangle the underlying (neurobiological) processes.

## Data Availability

The code used to generate the results of this study is available at https://github.com/Schrantee-lab. The data that support the findings of this study are not openly available due to privacy restrictions and are available from the corresponding author on reasonable request and execution of a data use agreement.

## Acknowledgements and disclosures

This study was funded by the Dutch non-profit organizations Kiddy GoodPills and Suffigium. The ePOD-MPH RCT (baseline data) was funded by a personal research grant awarded to LR by the Academic Medical Center, University of Amsterdam, and 11.32050.26ERA-NET PRIOMEDCHILD FP 6 (EU).

## Author contributions

Conceptualization: ZP, LR, HMG, AS. Formal analysis: ZP. Funding acquisition: LR. Investigation: AK, MAB, AS. Methodology: HJMM. Resources: LR. Supervision: LR, HMG, AS. Writing - original draft: ZP, LR, AS. Writing - review & editing: ZP, LR, HJJM, AK, MAB, HMG, AS.

A preprint version of this manuscript is available at https://doi.org/10.1101/2024.12.30.24319766. The baseline data have been included in previous publications about the randomized controlled trial [19, 32]. However, the data from the 4-year follow-up have not been published previously.

We would like to thank all participants and their parents for their contribution to this study and all students that helped with collection and analysis of the data.

The authors report no biomedical financial interests or potential conflicts of interests.

