## Supplementary Materials for "Association between long-term stimulant treatment and the functional brain response to methylphenidate in adolescents and adults with attention-deficit/hyperactivity disorder"

### SUPPLEMENTARY METHODS

#### Stimulant medication use

During the ePOD-methylphenidate RCT, the treating physician prescribed short-acting methylphenidate under double-blind clinical guidance (reduction of ADHD symptoms) following Dutch treatment guidelines. Exposure duration was 4 months for participants who received methylphenidate, and 0 months for participants who received placebo.

Stimulant medication use between ePOD-methylphenidate RCT end and 4-year follow-up assessment was calculated based on prescription information obtained from participants' pharmacies. Methylphenidate- and dexamphetamine-based formulations were considered as stimulant medication as treatment for ADHD, and cumulative dose was converted to methylphenidate-equivalents [1]. Exposure duration to stimulant medication was determined by calculating the time between the start date and end date of medication use, with a 30-day permissible gap to allow for commonly occurring "medication holidays", and rounding to months. The age at start of medication use was determined based on the date of the baseline assessment for participants who received methylphenidate during the ePOD-methylphenidate RCT, and date of first stimulant medication prescription in the pharmacy overview for participants who received placebo during the ePOD-methylphenidate RCT.

Participants with no stimulant medication use between baseline and 4-year follow-up assessment were considered stimulant treatment-naive, and cumulative dose and exposure duration were set to zero.

#### Adjusted MRI processing with the ExploreASL pipeline for six adolescents

Processing of the ASL data was adjusted for six adolescents. For two participants, background suppression was used for the ASL MR scans at baseline. For four adolescents, at 4-year follow-up the ASL MR data were exported as perfusion images instead of individual averages. For the four adolescents with differently exported ASL data at follow-up, a fixed value ($1E10^6$) was used as M0 instead of the mean of the control images. Next, for all six participants with adjusted ASL processing, the CBF images were divided by the mean M0 template of all other participants prior to calculation of regional and global CBFs. Finally, the mean CBFs of the adjusted participants were rescaled to the mean CBFs of the unadjusted participants.

#### Exploratory whole-brain analyses

<u>Relation between rCBF-response and stimulant medication use</u>

To assess the relation between whole-brain gray matter (GM) rCBF-response maps and stimulant medication use, we conducted linear regression analyses with Randomise [2] (5000 permutations). The rCBF-response to an acute methylphenidate-challenge was defined as *post-MPH - pre-MPH*. The significance level was set at $P_{FWE}$<.05, corrected for multiple comparisons using the threshold-free cluster enhancement (TFCE) option. Separate models were used per age group (adolescents, adults), medication use variable (cumulative dose, exposure duration), and visit (baseline, follow-up; we also tested the change between baseline and follow-up). Change in motion was included as a covariate in all models. At follow-up, for the four participants with differently exported ASL data (please see section above) no motion values were available, and the median change in motion across all other participants was used.

Relation between rCBF-response and dopamine and noradrenaline receptor distributions

To identify which receptor systems may be mediating the observed response, we explored the association between the whole-brain GM rCBF-response to methylphenidate and the spatial distribution of dopamine and noradrenaline receptors, separately for adolescents and adults. Whole-brain rCBF-response t-stat maps were generated using One-Sample t-tests with Randomise [2] (5000 permutations). Receptor distribution maps were obtained from neuromaps [3], an open source toolbox accessing, transforming and analyzing structural and functional brain annotations. We included the following maps (Supplementary Figure 1): dopamine transporter (DAT [4]), noradrenaline transporter (NAT [5]), dopamine D1 receptor [6], dopamine D2/D3 receptor [7]. All maps were resampled to 2mm isotropic resolution and parcellated into 250 atlas regions, based on the Cammoun atlas available in neuromaps [8]. To quantify the association between the rCBF-response maps and various receptor distributions, we employed a spin-test [9], a statistical method that computes null models by randomly rotating data, which allows for the assessment of statistical significance between two spatial maps [10]. Spatial null distributions were generated for the pre vs post-methylphenidate contrast based on the whole-brain rCBF-response t-stat maps using the approach outlined by Burt et al. [11] applying default parameters from the brainSMASH software with 1000 permutations, and these were subsequently correlated with each of the parcellated receptor maps. Given the exploratory nature of these analyses, we report the uncorrected p-values for each association.

**Supplementary Figure 1. Receptor distribution maps obtained from neuromaps [3].** Receptor maps are unnormalized and were resampled to 1mm isotropic resolution for visualization purposes. DAT = dopamine transporter; NAT = noradrenaline transporter; D1 = dopamine D1 receptor; D2/3 = dopamine D2/D3 receptor.

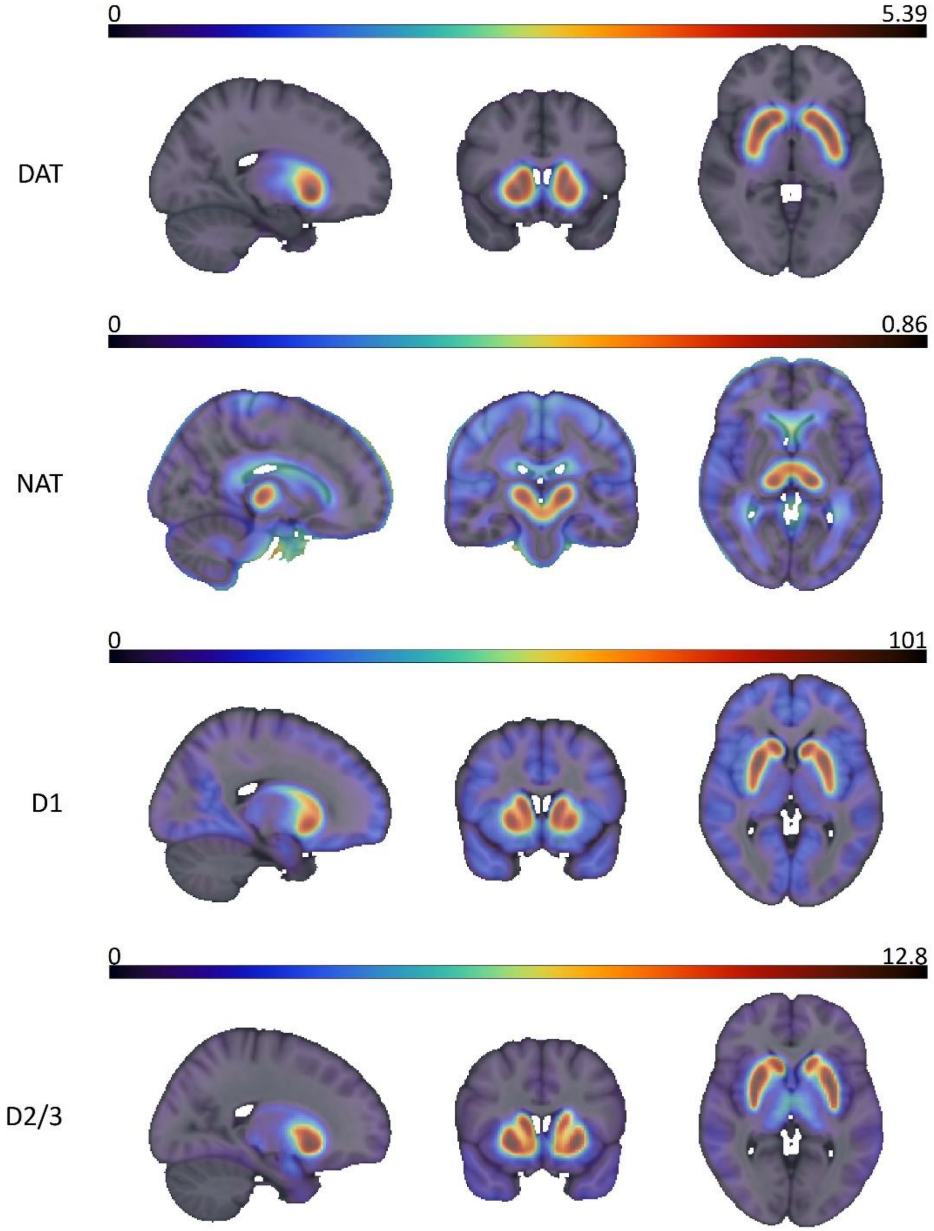

**Supplementary Table 1. Brainnetome parcellation numbers per region of interest (ROI).** ACC = anterior cingulate cortex; BNA = Brainnetome Atlas; mPFC = medial prefrontal cortex.

| Image | ROI | BNA parcellation numbers |
|---|---|---|
| 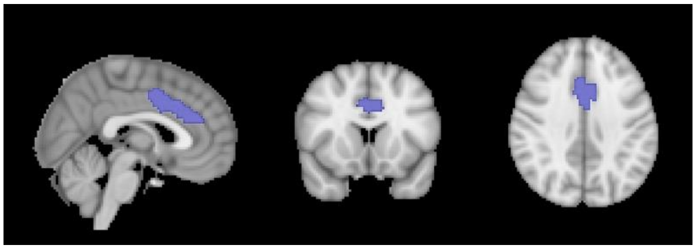 | **ACC** | **179, 180, 183, 184** |
| 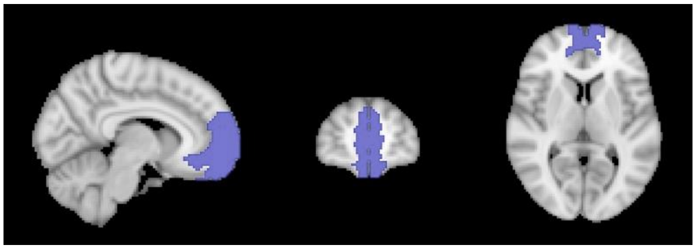 | **mPFC** | **13, 14, 41, 42, 47, 48** |
| 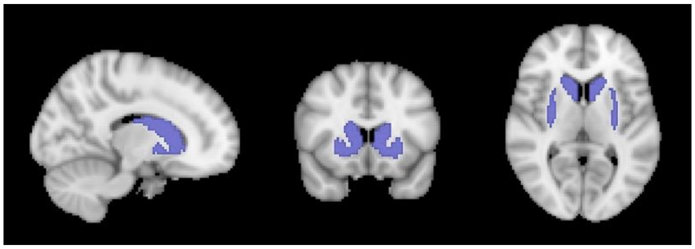 | **Striatum** | **219, 220, 225-230** |
| 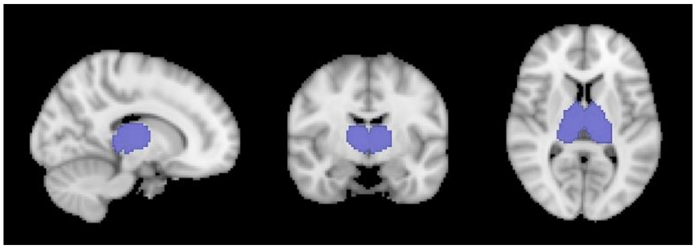 | **Thalamus** | **231-246** |

### SUPPLEMENTARY RESULTS

**Supplementary Table 2. Baseline characteristics of participants that did and did not participate in the 4-year follow-up assessment.** Data are presented as mean (standard deviation) or fraction (methylphenidate/placebo). Comparisons were made prior to data processing and analysis.

| | **Adolescents** | | | **Adults** | | |
|---|---|---|---|---|---|---|
| | Return = yes<br>n = 33 | Return = no<br>n = 17 | Statistics[a] | Return = yes<br>n = 25 | Return = no<br>n = 24 | Statistics[a] |
| Age<br>(years, *mean (SD)*) | 11.2 (0.9) | 11.6 (0.8) | t(48) = 1.49, P = .14 | 29.8 (5.0) | 27.3 (3.8) | t(47) = -1.98, P = .053 |
| IQ[b]<br>*(mean (SD)*) | 105.1 (17.7) | 102.3 (19.6) | t(46) = -0.50, P = .62 | 106.7 (4.9) | 109.0 (9.5) | t(43) = 1.05, P = .30 |
| ADHD-inattentive symptom severity[c]<br>*(mean (SD)*) | 22.60 (3.43) | 21.31 (2.98) | t(47) = -1.29, P = .20 | | | |
| ADHD-hyperactive/impulsive symptom severity[c]<br>*(mean (SD)*) | 15.85 (5.51) | 14.81 (6.50) | t(47) = -0.58, P = .56 | | | |
| ADHD symptom severity[d]<br>*(mean (SD)*) | | | | 33.00 (9.71) | 32.38 (9.83) | t(43) = -0.21, P = .83 |
| Anxiety symptoms[e]<br>*(mean (SD)*) | 26.03 (16.58) | 30.71 (17.50) | t(48) = 0.93, P = .36 | 6.84 (6.61) | 10.64 (7.03) | t(45) = 1.91, P = .06 |
| Depressive symptoms[f]<br>*(mean (SD)*) | 8.31 (4.75) | 8.29 (3.85) | t(47) = -0.01, P = .99 | 6.21 (4.90) | 8.18 (6.68) | t(44) = 1.15, P= .26 |
| ePOD-MPH RCT treatment group<br>(*methylphenidate/placebo)* | 12/21 | 13/4 | $X^2$(1) = 5.70, P = **.02** | 12/13 | 13/11 | $X^2$(1) = 0.02, P = .88 |

ADHD=attention-deficit/hyperactivity disorder, IQ=intelligence quotient. [a] Two-sample t-test or Chi-squared test. [b] For adolescents: subtest Wechsler Intelligence Scale for Children (WISC); for adults: National Adult Reading Test (NART, Dutch translation). [c] Inattentive and hyperactive/impulsive subscales of the Disruptive Behavior Disorder Rating-Scale (DBD-RS). [d] ADHD-Rating Scale (ADHD-RS) total score. [e] For adolescents: Screen for Child Anxiety Related Disorders (SCARED); for adults: Beck Anxiety Inventory (BAI). [f] For adolescents: Child Depression Inventory (CDI); for adults: Beck Depression Inventory (BDI).

In both adolescents and adults, participants that did and did not participate in the 4-year follow-up assessment did not differ in baseline characteristics (age, IQ, ADHD symptom severity, and symptoms of depression and anxiety). In adolescents, the proportion of ePOD-methylphenidate RCT groups (methylphenidate/placebo) differed between the participants who participated in the 4-year follow-up assessment and those who did not (X2(1)=5.70, P=.017; methylphenidate/placebo: 21/12 in return group, 4/13 in non-return group).

**Supplementary Table 3. Linear mixed effect model results for the models with treatment group during the trial (methylphenidate, placebo).** Bold values represent significant effects after multiple comparison correction (FDR=5%, 4 comparisons). The variables mean or change in motion, MR scanner at baseline, baseline age, and scan interval were included as covariates, but were excluded from the table for readability, except for motion, which showed a significant effect in one model. ACC=anterior cingulate cortex; mPFC=medial prefrontal cortex; $P_{FDR}$=P-value after FDR multiple comparison correction; RCT=randomized controlled trial.

| **A) Three-way linear mixed effects model with pre-MPH rCBF** | **ACC** | **mPFC** | **Striatum** | **Thalamus** |
|---|---|---|---|---|
| Age | F(1,103)=5.9, P=.017, $P_{FDR}$=.034 | F(1,62)=2.2, P=.14, $P_{FDR}$=.19 | F(1,61)=0.1, P=.74, $P_{FDR}$=.74 | **F(1,59)=17.4, P<.001, $P_{FDR}$<.001** |
| Visit | F(1,103)=0.1, P=.77, $P_{FDR}$=.77 | **F(1,56)=15.3, P<.001, $P_{FDR}$=.001** | F(1,51)=1.0, P=.32, $P_{FDR}$=.43 | **F(1,52)=10.8, P=.002, $P_{FDR}$=.004** |
| RCT treatment group | F(1,103)=0.1, P=.80, $P_{FDR}$=.80 | F(1,57)=1.0, P=.33, $P_{FDR}$=.80 | F(1,54)=0.1, P=.76, $P_{FDR}$=.80 | F(1,53)=0.2, P=.68, $P_{FDR}$=.80 |
| Age * Visit | F(1,103)<.01, P=.97, $P_{FDR}$=.97 | F(1,56)=0.6, P=.45, $P_{FDR}$=.97 | F(1,51)=0.1, P=.81, $P_{FDR}$=.97 | F(1,52)=0.1, P=.81, $P_{FDR}$=.97 |
| Age * RCT treatment group | F(1,103)=1.8, P=.19, $P_{FDR}$=.75 | F(1,56)=0.1, P=.74, $P_{FDR}$=.93 | F(1,54)<0.01, P=.93, $P_{FDR}$=.93 | F(1,53)=0.04, P=.85, $P_{FDR}$=.93 |
| Visit * RCT treatment group | F(1,103)=2.8, P=.10, $P_{FDR}$=.39 | F(1,56)=1.0, P=.32, $P_{FDR}$=.63 | F(1,52)=0.2, P=.65, $P_{FDR}$=.86 | F(1,52)=0.03, P=.86, $P_{FDR}$=.86 |
| Age * Visit * RCT treatment group | F(1,103)=0.3, P=.58, $P_{FDR}$=.77 | F(1,56)=1.4, P=.25, $P_{FDR}$=.77 | F(1,53)=0.5, P=.47, $P_{FDR}$=.77 | F(1,53)=0.03, P=.86, $P_{FDR}$=.86 |
| Mean Motion | F(1,103)=0.02, P=.90, $P_{FDR}$=.90 | F(1,99)=0.7, P=.39, $P_{FDR}$=.52 | F(1,103)=1.5, P=.22, $P_{FDR}$=.45 | **F(1,101)=, P=.012, $P_{FDR}$=.049** |
| **B) Three-way linear mixed effects model with rCBF-response to a MPH-challenge** | | | | |
| Age | F(1,98)=0.03, P=.87, $P_{FDR}$=.87 | F(1,54)=1.8, P=.19, $P_{FDR}$=.53 | F(1,52)=1.3, P=.26, $P_{FDR}$=.53 | F(1,41)=0.2, P=.67, $P_{FDR}$=.87 |
| Visit | F(1,98)=0.02, P=.89, $P_{FDR}$=.89 | F(1,49)=1.5, P=.22, $P_{FDR}$=.68 | F(1,51)=0.8, P=.78, $P_{FDR}$=.89 | F(1,40)=0.9, P=.34, $P_{FDR}$=.68 |
| RCT treatment group | F(1,98)=1.0, P=.33, $P_{FDR}$=.59 | F(1,50)=0.3, P=.59, $P_{FDR}$=.59 | F(1,50)=2.0, P=.16, $P_{FDR}$=.59 | F(1,39)=0.4, P=.52, $P_{FDR}$=.59 |
| Age * Visit | F(1,98)=1.2, P=.28, $P_{FDR}$=.56 | F(1,47)=5.9, P=.019, $P_{FDR}$=.075 | F(1,50)<0.01, P=.99, $P_{FDR}$=.99 | F(1,38)=0.5, P=.50, $P_{FDR}$=.66 |
| Age * RCT treatment group | F(1,98)=0.4, P=.51, $P_{FDR}$=.77 | F(1,51)=0.5, P=.50, $P_{FDR}$=.77 | F(1,51)=0.1, P=.70, $P_{FDR}$=.77 | F(1,39)=0.1, P=.77, $P_{FDR}$=.77 |
| Visit * RCT treatment group | F(1,98)=0.8, P=.38, $P_{FDR}$=.76 | F(1,48)=0.02, P=.90, $P_{FDR}$=.90 | F(1,50)=1.4, P=.24, $P_{FDR}$=.76 | F(1,39)=0.2, P=.70, $P_{FDR}$=.90 |
| Age * Visit * RCT treatment group | F(1,98)=1.0, P=.32, $P_{FDR}$=.45 | F(1,46)=1.2, P=.28, $P_{FDR}$=.45 | F(1,49)=0.1, P=.81, $P_{FDR}$=.81 | F(1,37)=0.9, P=.34, $P_{FDR}$=.45 |
| Change in Motion | F(1,98)=1.8, P=.18, $P_{FDR}$=.36 | F(1,97)=5.6, P=.020, $P_{FDR}$=.082 | F(1,85)=0.2, P=.64, $P_{FDR}$=.66 | F(1,83)=0.2, P=.66, $P_{FDR}$=.66 |

**Supplementary Table 4. Cerebral blood flow (CBF) values used to calculate relative CBF (rCBF) for analysis.** Data are presented as mean (standard deviation; SD) and n of the CBF values (mL/min/100g) that were estimated using the ExploreASL pipeline. Bold values represent significant differences. Six participants (two baseline, four follow-up) were excluded, since changes were made to the ExploreASL pipeline (see Supplementary Methods). ACC=anterior cingulate cortex; GM=grey matter; mPFC=medial prefrontal cortex; MPH=methylphenidate.

| | | **Adolescents** | | | | | **Adults** | | | | |
|---|---|---|---|---|---|---|---|---|---|---|---|
| | | pre-MPH | n | post-MPH | n | Statistics[a] | pre-MPH | n | post-MPH | n | Statistics[a] |
| *Baseline* | | | | | | | | | | | |
| | ACC | 106.6 (15.4) | 28 | 98.1 (11.9) | 27 | t(24)=1.5, P=.14 | 88.5 (11.0) | 23 | 82.7 (11.0) | 24 | t(22)=4.4, **P<.001** |
| | mPFC | 93.7 (10.5) | 29 | 92.9 (7.1) | 27 | t(25)=0.3, P=.80 | 75.8 (10.7) | 23 | 74.0 (10.0) | 24 | t(22)=1.8, P=.08 |
| | Striatum | 82.1 (12.3) | 28 | 78.8 (7.8) | 27 | t(24)=0.7, P=.48 | 66.9 (9.9) | 23 | 67.4 (9.5) | 24 | t(22)=-0.5, P=.62 |
| | Thalamus | 70.2 (11.4) | 29 | 67.0 (8.8) | 27 | t(25)=0.9, P=.35 | 64.8 (9.3) | 23 | 64.8 (8.5) | 24 | t(22)=0.1, P=.89 |
| | Total GM | 84.4 (9.1) | 29 | 80.0 (7.5) | 27 | t(25)=2.2, **P=.04** | 67.1 (9.0) | 23 | 65.2 (8.8) | 24 | t(22)=2.3, **P=.03** |
| *Follow-up* | | | | | | | | | | | |
| | ACC | 90.4 (15.1) | 26 | 86.9 (10.4) | 26 | t(23)=2.5, **P=.02** | 84.5 (13.3) | 24 | 80.4 (12.8) | 24 | t(23)=3.2, **P=.004** |
| | mPFC | 82.7 (14.0) | 26 | 81.9 (12.8) | 26 | t(23)=1.4, P=.19 | 74.5 (10.9) | 24 | 73.4 (11.4) | 24 | t(23)=1.0, P=.33 |
| | Striatum | 71.4 (10.0) | 26 | 71.7 (8.9) | 26 | t(23)=0.7, P=.49 | 64.2 (10.3) | 24 | 64.4 (10.6) | 24 | t(23)=-0.2, P=.84 |
| | Thalamus | 56.8 (11.1) | 26 | 59.2 (10.5) | 26 | t(23)=-1.5, P=.14 | 58.1 (10.6) | 24 | 58.5 (10.7) | 24 | t(23)=-0.4, P=.72 |
| | Total GM | 71.5 (12.2) | 26 | 71.2 (10.7) | 26 | t(23)=0.9, P=.38 | 63.7 (9.5) | 24 | 62.0 (9.7) | 24 | t(23)=1.6, P=.12 |

[a] Paired-samples t-test.

Resting CBF (i.e. before the methylphenidate-challenge) reduced between baseline and follow-up in all regions of interest in adolescents (ACC: t(21)=3.7, P=.001; mPFC: t(22)=3.7, P=.001; striatum: t(21)=3.4, P=.002; thalamus: t(22)=5.1, P<.001; total GM: t(22)=4.6, P<.001), but not in adults (ACC: t(22)=1.5, P=.14; mPFC: t(22)=0.5, P=.61; striatum: t(22)=1.3, P=.22; thalamus: t(22)=3.2, P=.004; total GM: t(22)=1.8, P=.08).

**Supplementary Table 5. Pearson correlations between ADHD symptom severity and rCBF for regions of interest (ROIs) showing significant age-dependent medication effects.** The column 'change in symptoms' shows the correlation with baseline rCBF. ADHD=attention-deficit/hyperactivity disorder; ADHD-RS=ADHD Rating Scale; BL=baseline; DBD-RS=Disruptive behavior disorder rating scale; FU=follow-up; mPFC=medial prefrontal cortex; rCBF=relative cerebral blood flow.

| | **Adolescents** | | | **Adults** | | |
|---|---|---|---|---|---|---|
| | **BL** | **FU** | **Change in symptoms** | **BL** | **FU** | **Change in symptoms** |
| *Thalamic resting rCBF* | | | | | | |
| DBD-RS inattentive | r=-.02, P=.93 | r=.01, P=.98 | r=-.22, P=.26 | | | |
| DBD-RS hyperactive/impulsive | r=-.11, P=.57 | r=-.01, P=.97 | r=.05, P=.80 | | | |
| ADHD-RS | | | | r=.02, P=.94 | r=.34, P=.16 | r=.30, P=.22 |
| *mPFC rCBF-response to a methylphenidate-challenge* | | | | | | |
| DBD-RS inattentive | r=-.21, P=.28 | r=.28, P=.17 | r=-.05, P=.81 | | | |
| DBD-RS hyperactive/impulsive | r=-.05, P=.79 | r=.30, P=.14 | r=.11, P=.60 | | | |
| ADHD-RS | | | | r=.34, P=.13 | r=-.06, P=.79 | r=-.27, P=.27 |

In both age groups, we found no associations between change in ADHD symptom severity and change in thalamic resting rCBF (adolescents: inattentive: r=.27, P=19; hyperactive/impulsive: r=-.08, P=.70; adults: r=.25, P=.30). The association between change in ADHD symptom severity and change in rCBF was not tested for the mPFC rCBF-response, as the rCBF-response in this region did not significantly change between baseline and follow-up.

**Supplementary Table 6. Heart rate before (pre-MPH) and after (post-MPH) a single-dose methylphenidate-challenge.** Data are presented as mean (standard deviation; SD). Heart rate increased after the methylphenidate-challenge. BL=baseline; FU=follow-up; MPH=methylphenidate.

| | **Adolescents** | | | **Adults** | | |
|---|---|---|---|---|---|---|
| | pre-MPH | post-MPH | Statistics[a] | pre-MPH | post-MPH | Statistics[a] |
| BL | 69.6 (8.2) | 86.5 (9.9) | t(20)=-11.1, P<.001 | 62.9 (9.3) | 80.7 (15.2) | t(22)=-6.9, P<.001 |
| FU | 63.6 (2.1) | 85.9 (8.6) | t(16)=-11.0, P<.001 | 62.9 (13.2) | 80.8 (12.5) | t(18)=-6.4, P<.001 |

[a] Paired-samples t-test.

**Supplementary Table 7. Pearson correlations between heart rate and rCBF-response to a single-dose methylphenidate-challenge.** We found no significant associations between heart rate and rCBF-response. ACC=anterior cingulate cortex; BL=baseline; FU=follow-up; mPFC=medial prefrontal cortex; $P_{FDR}$=P-value after FDR multiple comparison correction; rCBF=relative cerebral blood flow.

| **rCBF-response to a methylphenidate-challenge** | **Adolescents** | | **Adults** | |
|---|---|---|---|---|
| | **BL** | **FU** | **BL** | **FU** |
| ACC | r=.22, P=.36, $P_{FDR}$=.48 | r=-.15, P=.57, $P_{FDR}$=.68 | r=-.38, P=.07, $P_{FDR}$=.20 | r=.04, P=.88, $P_{FDR}$=.88 |
| mPFC | r=-.21, P=.36, $P_{FDR}$=.48 | r=-.11, P=.68, $P_{FDR}$=.68 | r=.11, P=.63, $P_{FDR}$=.63 | r=.26, P=.29, $P_{FDR}$=.88 |
| Striatum | r=.01, P=.97, $P_{FDR}$=.97 | r=-.20, P=.43, $P_{FDR}$=.68 | r=-.35, P=.10, $P_{FDR}$=.20 | r=-.05, P=.85, $P_{FDR}$=.88 |
| Thalamus | r=.24, P=.30, $P_{FDR}$=.48 | r=.31, P=.23, $P_{FDR}$=.68 | r=.14, P=.53, $P_{FDR}$=.63 | r=-.07, P=.78, $P_{FDR}$=.88 |

**Supplementary Table 8. Motion (mm) before (pre-MPH) and after (post-MPH) a single-dose methylphenidate-challenge.** Data are presented as median (interquartile range). Motion was significantly higher in adolescents versus adults, and reduced after the methylphenidate-challenge in adolescents at follow-up only. BL=baseline; FU=follow-up; MPH=methylphenidate.

| | **Adolescents** | | | **Adults** | | | **Adolescents vs Adults**[a] | |
|---|---|---|---|---|---|---|---|---|
| | pre-MPH | post-MPH | Statistics[a] | pre-MPH | post-MPH | Statistics[a] | pre-MPH | post-MPH |
| BL | 0.20 (0.15-0.58) | 0.15 (0.11-0.29) | W=561, P=.10 | 0.07 (0.06-0.09) | 0.07 (0.05-0.08) | W=288, P=.81 | W=661, P<.001 | W=647, P<.001 |
| FU | 0.15 (0.11-0.30) | 0.11 (0.08-0.15) | W=492, P=.004 | 0.06 (0.04-0.08) | 0.05 (0.04-0.07) | W=337, P=.32 | W=573, P<.001 | W=547, P<.001 |

[a]Mann-Whitney U test.

**Supplementary Table 9. Spearman correlations between motion and rCBF.** After correction for multiple comparisons, we found no significant associations between motion and rCBF. ACC=anterior cingulate cortex; BL=baseline; FU=follow-up; mPFC=medial prefrontal cortex; $P_{FDR}$=P-value after FDR multiple comparison correction; rCBF=relative cerebral blood flow.

| **A) Resting rCBF** | **Adolescents** | | **Adults** | |
|---|---|---|---|---|
| | **BL** | **FU** | **BL** | **FU** |
| ACC | r=-.07, P=.72, $P_{FDR}$=.72 | r=-.11, P=.59, $P_{FDR}$=.90 | r=.22, P=.32, $P_{FDR}$=.63 | r=-.07, P=.76, $P_{FDR}$=.76 |
| mPFC | r=-.34, P=.06, $P_{FDR}$=.12 | r=-.03, P=.90, $P_{FDR}$=.90 | r=-.25, P=.24, $P_{FDR}$=.63 | r=-.29, P=.17, $P_{FDR}$=.63 |
| Striatum | r=-.23, P=.22, $P_{FDR}$=.30 | r=-.06, P=.78, $P_{FDR}$=.90 | r=-.05, P=.83, $P_{FDR}$=.91 | r=-.16, P=.46, $P_{FDR}$=.63 |
| Thalamus | r=-.40, P=.03, $P_{FDR}$=.11 | r=-.23, P=.25, $P_{FDR}$=.90 | r=.03, P=.91, $P_{FDR}$=.91 | r=-.15, P=.47, $P_{FDR}$=.63 |
| **B) rCBF-response to a methylphenidate-challenge** | | | | |
| | **BL** | **FU** | **BL** | **FU** |
| ACC | r=-.17, P=.38, $P_{FDR}$=.51 | r=-.32, P=.12, $P_{FDR}$=.50 | r=-.24, P=.26, $P_{FDR}$=.99 | r=.28, P=.19, $P_{FDR}$=.59 |
| mPFC | r=.34, P=.08, $P_{FDR}$=.26 | r=-.07, P=.74, $P_{FDR}$=.74 | r=-.05, P=.81, $P_{FDR}$=.99 | r=-.16, P=.44, $P_{FDR}$=.59 |
| Striatum | r=-.12, P=.56, $P_{FDR}$=.56 | r=.08, P=.70, $P_{FDR}$=.74 | r=-.09, P=.69, $P_{FDR}$=.99 | r=.05, P=.80, $P_{FDR}$=.80 |
| Thalamus | r=-.29, P=.13, $P_{FDR}$=.26 | r=.09, P=.69, $P_{FDR}$=.74 | r=-<.01, P=.99, $P_{FDR}$=.99 | r=.19, P=.38, $P_{FDR}$=.59 |
